# Are fewer cases of diabetes mellitus diagnosed in the months after SARS-CoV-2 infection?

**DOI:** 10.1101/2022.12.02.22283029

**Authors:** Neha V Reddy, Hsin-Chieh Yeh, Jena S Tronieri, Til Stürmer, John B Buse, Jane E Reusch, Steven G Johnson, Rachel Wong, Richard Moffitt, Kenneth J Wilkins, Jeremy Harper, Carolyn T Bramante, the N3C and RECOVER Consortiums

**Author notes:** **Corresponding Author:** Neha V. Reddy, ScB. Authors contributed equally (Jeremy Harper, Carolyn T Bramante).

## Abstract

Long-term sequelae of severe acute respiratory coronavirus-2 (SARS-CoV-2) infection may include an increased incidence of diabetes. Our objective was to describe the temporal relationship between new diagnoses of diabetes mellitus and SARS-CoV-2 infection in a nationally representative database. There appears to be a sharp increase in diabetes diagnoses in the 30 days surrounding SARS-CoV-2 infection, followed by a decrease in new diagnoses in the post-acute period, up to 360 days after infection. These results underscore the need for further investigation, as understanding the timing of new diabetes onset after COVID-19 has implications regarding potential etiology and screening and treatment strategies.

## Introduction

The relationship between severe acute respiratory coronavirus 2 (SARS-CoV-2), the Covid-19 pandemic, and diabetes mellitus (DM) is the subject of active investigation (1-2). A growing body of evidence suggests a possible increased incidence of new-onset DM after infection with SARS-CoV-2 (3-5). The mechanism by which SARS-CoV-2 infection increases the risk of developing DM is not fully understood. Hypotheses include stress hyperglycemia related to acute illness, direct effect of the virus on pancreatic vasculature or beta cells, changes in innate immunity, and iatrogenic causes (6-8).

However, risk factors for developing diabetes likely increased for the population, regardless of infection with SARS-CoV-2, as rates of overweight and obesity increased at the population level (9-11). In addition, physical activity declined globally during the pandemic and has not recovered (12). Stress related to the pandemic has increased endogenous cortisol which may be risk factor for DM (10, 13-14). It is currently unknown if rates of incident DM are higher after infection with SARS-CoV-2 than after infection with similar non-Covid-19 viruses (3-5).

We sought to characterize the temporal relationship between new diabetes diagnoses relative to SARS-CoV-2 infection in individuals who had SARS-CoV-2 and were also diagnosed with DM within 6 months of the start of the pandemic. We undertook this analysis in a large, nationally representative database in the U.S., the National Covid Cohort Collaborative (N3C).

## Methods

This is a nation-wide cross-sectional analysis to display the temporal relationship between new onset diabetes relative to SARS-CoV-2 infection in the N3C database. The N3C database aggregates data from electronic health records (EHR) from more than 70 institutions nationally to accelerate research efforts on the evolving Covid-19 pandemic (15).

The study sample included all persons with SARS-CoV-2 and with type 2 DM, from March 2020 to February 2022 in the N3C Database. To reduce ascertainment and selection bias, the sample was restricted to patients who had at least one outpatient clinical encounter at least six months prior to their SARS-CoV-2 infection, and at least 6 months of follow-up data. Because the earliest data in the N3C database is from 2018, everyone in the sample had engaged with the healthcare system at least once between 6 and 21 months prior to their SARS-CoV-2 infection.

SARS-CoV-2 infection is defined by either the ICD-10 code or laboratory results confirmed in a clinical setting. The date of SARS-CoV-2 infection is the index date and includes the 7 days prior to the index date to account for labs that resulted after the initiation of a clinical encounter for suspected Covid-19.

For this manuscript, type 2 diabetes is defined by ICD code using ICD lists reviewed for completeness and accuracy by two clinicians (16). New DM is defined as the earliest problem list code for DM, in persons who did not have an ICD code for DM in the EHR before September 2019. Because the sample is restricted to persons who had at least 6 months of EHR data before an infection with SARS-CoV-2, the earliest DM diagnoses in the sample would be September 2019. All problem list items in N3C are mapped to SNOMED, and 523 SNOMED codes were identified by clinicians as a diagnosis of DM. The earliest DM diagnosis date was subtracted from the index date per subject to determine a specific number of days to bucket them per 30-day time period.

Two analyses of DM incidence were conducted among persons in the N3C who had SARS-CoV-2 and an ICD code for DM, but who did not have an ICD code for DM before September 2019. Both analyses are presented by calendar month of the pandemic. Analyses were conducted in the secure computing environment using Palantir and R statistical software (R Foundation for Statistical Computing, Vienna, Austria).

## Results

Table 1 presents the demographic characteristics of new cases of DM in persons with SARS-CoV-2 infection in the N3C, by each month of the pandemic. Figures 1 and 2 present the temporal relationship between SARS-CoV-2 infection and new diagnoses of DM. Figure 1 shows the proportion of individuals with DM that were diagnosed with DM since September 2019; Figure 2 shows the number of new DM cases relative to SARS-CoV-2 infection. DM diagnosed in the 7 days prior SARS-CoV-2 infection could indicate that the DM and SARS-CoV-2 were diagnosed in the same healthcare encounter, due to lag times in SARS-CoV-2 results.

**Table 1:** Demographic characteristics of all patients in the N3C Data Enclave with SARS-CoV-2 infection from March 2020 to June 2022 and a clinical encounter 6 months prior to infection. Demographic characteristics are reported for all individuals in the sample who received a new diabetes diagnosis in the six months prior to 360 days after SARS-CoV-2 infection, across each month of the pandemic.

| Day relative to Calendar Month | Person count | Age | Female |  | Race/Ethnicity, n (%) |  |  |  |  |  |  |  |  |  |
| --- | --- | --- | --- | --- | --- | --- | --- | --- | --- | --- | --- | --- | --- | --- |
|  |  |  | n (%) | Age | Asian/<br>Pacific<br>Islander | Age | Black | Age | White | Age | Hispanic/Latinx | Age | Un-known | Age |
| 3-Mar-20 | 21988 | 54 | 12070 (55) | 45 | 921 (4) | 43 | 5648 (26) | 45 | 10415 (47) | 46 | 3313 (15) | 42 | 99 (0.4) | 43 |
| 4-Apr-20 | 63018 | 54 | 37102 (59) | 49 | 2413 (4) | 48 | 15689 (25) | 48 | 28650 (45) | 50 | 13068 (21) | 45 | 107 (0.2) | 43 |
| 5-May-20 | 49149 | 51 | 29598 (60) | 47 | 1834 (4) | 45 | 10847 (22) | 46 | 24188 (49) | 49 | 11629 (24) | 43 | 59 (0.1) | 42 |
| 6-Jun-20 | 41683 | 48 | 25146 (60) | 49 | 1433 (3) | 48 | 7877 (19) | 48 | 23726 (57) | 51 | 9099 (22) | 46 | 139 (0.3) | 43 |
| 7-Jul-20 | 66264 | 46 | 40103 (61) | 45 | 1742 (3) | 41 | 12912 (19) | 42 | 40829 (62) | 47 | 12298 (19) | 41 | 253 (0.4) | 40 |
| 8-Aug-20 | 59857 | 46 | 36204 (60) | 48 | 1570 (3) | 45 | 9110 (15) | 47 | 39750 (66) | 50 | 9890 (17) | 43 | 155 (0.3) | 39 |
| 9-Sep-20 | 55507 | 47 | 32691 (59) | 46 | 1445 (3) | 44 | 7012 (13) | 43 | 39216 (71) | 49 | 7619 (14) | 41 | 116 (0.2) | 45 |
| 10-Oct-20 | 106177 | 49 | 62061 (58) | 46 | 2534 (2) | 44 | 11501 (11) | 42 | 79630 (75) | 48 | 12127 (11) | 41 | 136 (0.1) | 42 |
| 11-Nov-20 | 244876 | 48 | 143786 (59) | 49 | 5927 (2) | 49 | 25786 (11) | 49 | 185952 (76) | 51 | 26086 (11) | 47 | 439 (0.2) | 47 |
| 12-Dec-20 | 253434 | 50 | 149547 (59) | 46 | 7091 (3) | 45 | 31757 (13) | 44 | 182328 (72) | 47 | 30440 (12) | 43 | 718 (0.3) | 39 |
| 13-Jan-21 | 177844 | 50 | 104217 (59) | 49 | 5216 (3) | 47 | 23838 (13) | 49 | 122835 (69) | 52 | 22905 (13) | 45 | 529 (0.3) | 46 |
| 14-Feb-21 | 72152 | 50 | 41647 (58) | 50 | 2358 (3) | 44 | 10348 (14) | 48 | 47011 (65) | 54 | 9178 (13) | 46 | 107 (0.2) | 43 |
| 15-Mar-21 | 59866 | 48 | 34893 (58) | 51 | 1940 (3) | 47 | 8930 (15) | 48 | 38545 (64) | 54 | 6820 (11) | 46 | 50 (0.1) | 43 |
| 16-Apr-21 | 63475 | 46 | 37704 (59) | 51 | 1838 (3) | 51 | 11917 (19) | 49 | 40070 (63) | 54 | 6632 (10) | 46 | 58 (0.1) | 48 |
| 17-May-21 | 37838 | 48 | 22835 (60) | 47 | 943 (2) | 46 | 7651 (20) | 44 | 23756 (63) | 49 | 3663 (10) | 42 | <20 | 49 |
| 18-Jun-21 | 19748 | 51 | 11747 (59) | 46 | 580 (3) | 48 | 3019 (15) | 41 | 13247 (67) | 49 | 1770 (9) | 41 | 23 (0.1) | 42 |
| 19-Jul-21 | 39539 | 47 | 23260 (59) | 47 | 903 (2) | 44 | 7142 (18) | 44 | 26810 (68) | 48 | 3196 (8) | 42 | 145 (0.4) | 41 |
| 20-Aug-21 | 126287 | 47 | 74135 (59) | 47 | 2273 (2) | 47 | 19468 (15) | 47 | 91961 (73) | 48 | 9395 (7) | 46 | 247 (0.2) | 43 |
| 21-Sep-21 | 120129 | 47 | 70806 (59) | 48 | 1910 (2) | 46 | 15700 (13) | 44 | 91260 (76) | 50 | 8214 (7) | 42 | 151 (0.1) | 43 |
| 22-Oct-21 | 75960 | 49 | 44359 (58) | 50 | 1143 (2) | 50 | 8060 (11) | 45 | 59436 (78) | 53 | 5046 (7) | 44 | 76 (0.1) | 39 |
| 23-Nov-21 | 97887 | 48 | 56396 (58) | 50 | 1563 (2) | 44 | 9111 (9) | 49 | 78025 (80) | 54 | 6224 (6) | 46 | 66 (0.1) | 44 |
| 24-Dec-21 | 224781 | 45 | 134429 (60) | 52 | 5517 (2) | 51 | 41095 (18) | 53 | 149546 (67) | 56 | 19257 (9) | 49 | 538 (0.2) | 52 |
| 25-Jan-22 | 424738 | 46 | 257183 (61) | 45 | 13960 (3) | 44 | 60657 (14) | 45 | 295523 (70) | 47 | 42728 (10) | 43 | 1323 (0.3) | 40 |
| 26-Feb-22 | 76582 | 51 | 46117 (60) | 50 | 2220 (3) | 48 | 7408 (10) | 49 | 57584 (75) | 53 | 6842 (9) | 44 | 227 (0.3) | 47 |
| 27-Mar-22 | 24794 | 52 | 14986 (60) | 45 | 964 (4) | 44 | 2264 (9) | 47 | 18210 (73) | 47 | 2188 (9) | 43 | 82 (0.3) | 43 |
| 28-Apr-22 | 35539 | 52 | 21736 (61) | 47 | 1905 (5) | 45 | 3279 (9) | 47 | 26350 (74) | 49 | 2640 (7) | 44 | 194 (0.6) | 42 |
| 29-May-22 | 54951 | 52 | 34055 (62) | 47 | 3136 (6) | 44 | 5706 (10) | 48 | 39729 (72) | 48 | 4570 (8) | 44 | 404 (0.8) | 39 |
| 30-Jun-22 | 31044 | 52 | 19280 (62) | 52 | 1570 (5) | 49 | 3815 (12) | 55 | 21914 (71) | 54 | 3430 (11) | 51 | 155 (0.5) | 53 |

**Figure 1:**
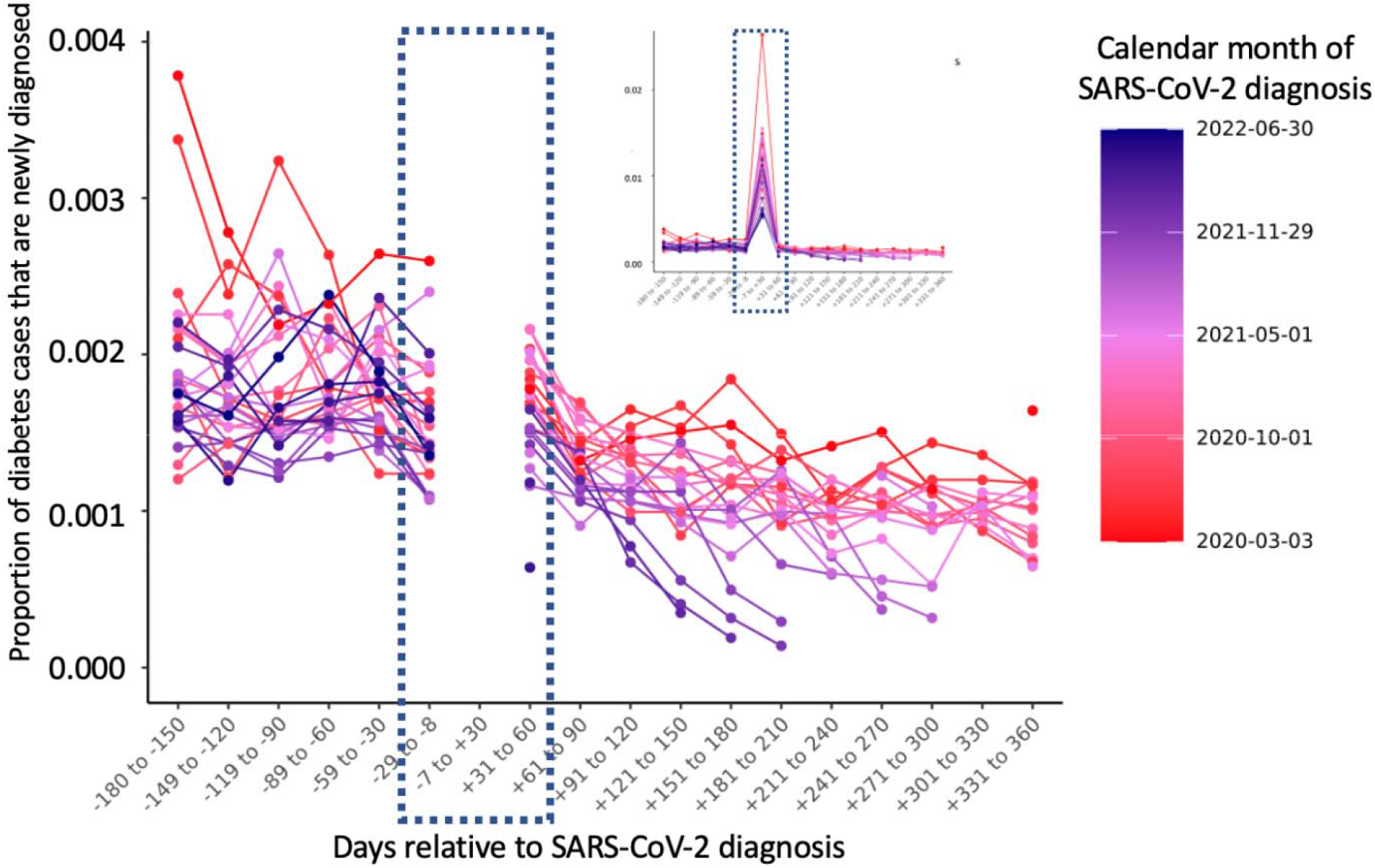
Among all persons with diabetes in the database, the proportion of diabetes cases that were diagnosed between Sept 2019 and February 2022, by 30-day periods relative to infection with SARS-CoV-2. This plot represents the proportion of diabetes cases in the N3C database that were diagnosed after September 2019 in persons who did not have a previous diagnosis of diabetes. Each line represents a calendar month of the pandemic. The Y axis is the proportion of all persons with SARS-CoV-2 and a DM diagnosis who received the ICD code for DM after September 2019, and the temporal relationship between the diagnoses between 180 days prior to 360 days after the SARS-Co-V-2 infection. The large peak between 8 days before and 30 days after is represented in an inset so that the top of that peak is visible without compressing the Y axis.

**Figure 2:**
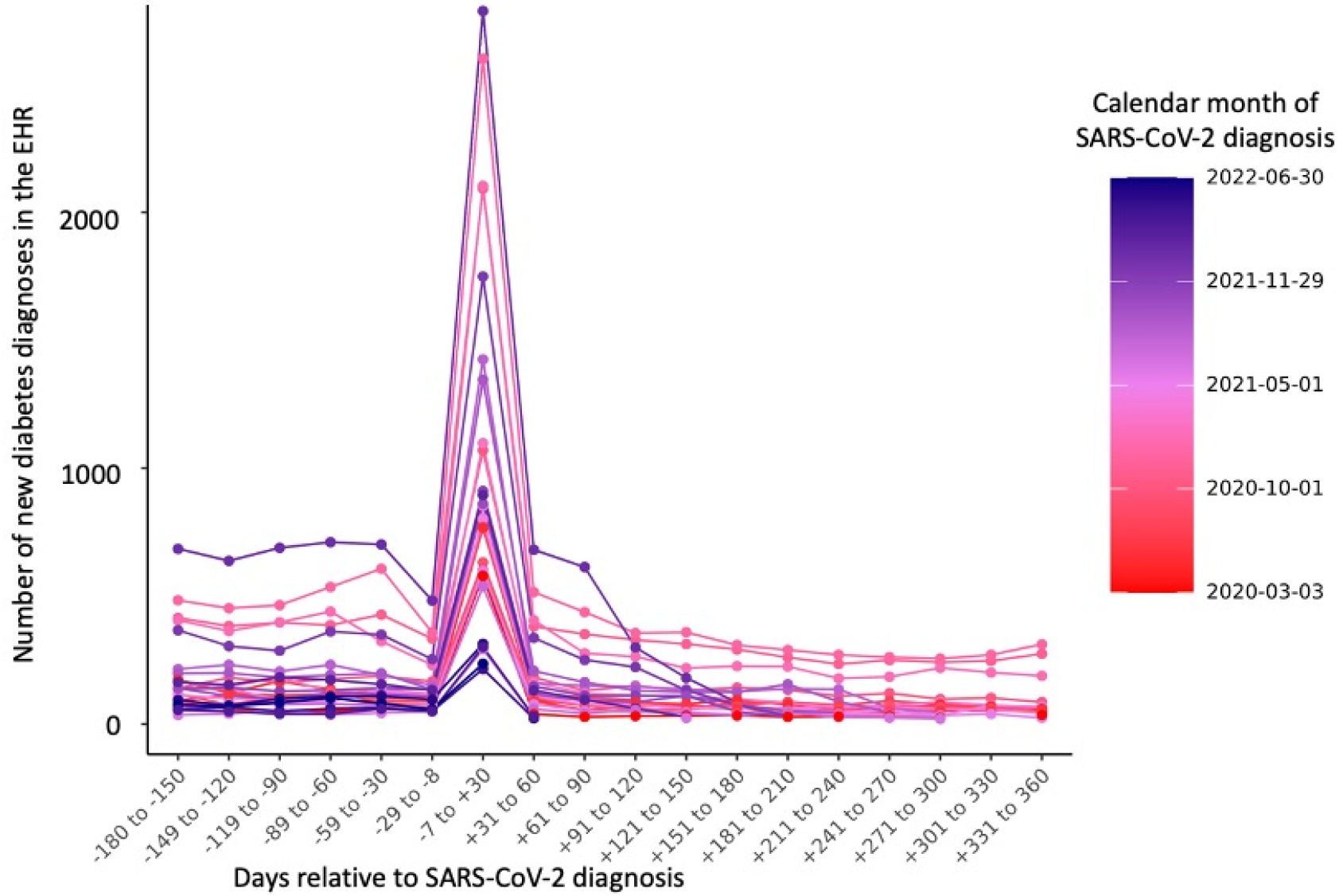
Number of new diabetes mellitus (DM) cases within 180 before and 360 days after SARS-CoV-2 infection, among individuals who have both EHR-recorded SARS-CoV-2 infection and ICD code for DM and did not have an ICD code for DM before September 2019. Each line represents a calendar month during the pandemic; the Y axis is the number of new DM cases in the N3C database in persons with a documented SARS-CoV-2 infection. The x-axis is temporal relationship between the diabetes diagnosis relative to each individual’s SARS-CoV-2 infection.

Both figures depict a sharp increase in DM diagnoses in the days immediately surrounding a positive SARS-CoV-2 result, from 7 days prior to 30 days after SARS-CoV-2 infection. The count of new DM diagnoses is lower in the months after SARS-CoV-2 infection than in the months before (Figure 2). The percentage of DM cases in the EHR that are newly diagnosed appears to be approximately 2% to 3% lower for each month after the index date (Figure 2). Supplementary Tables 1 and 2 contain the data that were used to make the figures.

## Discussion

This is a cross-sectional, population-level assessment of the temporal relationship between SARS-CoV-2 infection and new diagnoses of DM persons who have both conditions in the N3C database. We observed a sharp increase in new cases of DM within 7 days before to 30 days after SARS-CoV-2 infection, followed by a decrease in new DM diagnoses. It also appears that the number and proportion of new-onset DM may then be lower in the months after SARS-Co-V-2 infection than in the months preceding.

It is possible that the sharp peak in DM cases represents a marked increase of interaction with the healthcare system at the time of SARS-CoV-2 infection. Hemoglobin A1C is the most commonly used criteria for diagnosing diabetes. Because of the general requirement for confirmation of lab criteria for the diagnosis of diabetes and the asynchronous documentation of ICD codes at an encounter with the return of lab results, clinicians may not enter an ICD code for DM after just one episode of hyperglycemia, or two episodes of hyperglycemia in the same healthcare encounter. Thus, the spike of new DM cases immediately surrounding SARS-CoV-2 infections could represent individuals who were newly interacting with the healthcare system, having their hemoglobin A1C or glucose value tested, and receiving a diagnosis of DM based on the lab result.

A recent retrospective cohort study reported an increased risk of incident DM during the post-acute infection period in the US Department of Veterans Affairs (VA) database (5). The VA population has guaranteed access to healthcare, whereas the spike followed by a decrease in new-onset DM in the N3C population may reflect patients newly interacting with the healthcare system who are simply *diagnosed* with DM that they already had because of this interaction with healthcare. Further analysis from the VA cohorts may demonstrate a decreased incidence of new-onset DM when followed for a full year after infection.

Another potential explanation for a large peak in DM cases is that the physiologic stress of infection with SARS-CoV-2 does push high-risk individuals to develop DM. The decrease in new DM diagnoses in the months after SARS-CoV-2 infection could reflect the metabolic challenge of the infection revealing DM that would have presented later in the absence of SARS-CoV-2 infection. As such the SARS-CoV-2 infection caused individuals to develop diabetes earlier, or be diagnosed earlier, thereby decreasing the population at risk for DM in the months after SARS-CoV-2 infection.

Understanding the population at risk for diabetes is difficult when considering the effects of the pandemic on the population as a whole – increased stress, weight, central adiposity and decreased physical activity. Thus, the risk of developing diabetes has potentially increased regardless of infection with SARS-CoV-2. It is also possible that infection with SARS-CoV-2 does precipitate new diagnoses of DM in persons who would not have developed diabetes otherwise, and that with longer follow-up the slope of new DM diagnoses will go up in persons who have been infected with SARS-CoV-2.

This is not a causal analysis and should not be interpreted as such. Our analysis suggests that there is a spike in DM diagnoses immediately surrounding SARS-CoV-2 infection, followed by a decrease in new diagnoses. Such a pattern may be attributable to increased interaction with the healthcare system or the stress of the SARS-CoV-2 infection. Rigorous epidemiologic and mechanistic studies are needed to understand whether there are causal relationships between SARS-CoV-2 infection and the development of DM in the short- and long-term. Clinical trial cohorts may represent an opportunity for prospective, complete data for assessing the prevalence of new-onset DM in those infected with SARS-CoV-2. The main conclusion from this cross-sectional analysis of the temporal relationship between SARS-CoV-2 infection and DM is that the interplay between Covid-19, pandemic related lifestyle chances and DM is complex and must be studied carefully with an appropriate control cohort.

## Data Availability

All data produced in the present work are contained in the manuscript and supplemental materials.

## Author Contributions

J.H. and R.M. completed the data analysis. N.V.R., H.Y., J.S.T, T.S., J.B.B., J.E.R., S.G.J, R.W., K.J.W, and C.T.B. contributed to the scientific design, critical review, and revisions of the manuscript.

## Acknowledgements

The content is solely the responsibility of the authors and does not necessarily represent the official views of the RECOVER Program, the NIH or other funders.

The analyses described in this publication were conducted with data or tools accessed through the NCATS N3C Data Enclave Covid.cd2h.org/enclave and supported by CD2H - The National Covid Cohort Collaborative (N3C) IDeA CTR Collaboration 3U24TR002306-04S2 NCATS U24 TR002306. This research was possible because of the patients whose information is included within the data from participating organizations (Covid.cd2h.org/dtas) and the organizations and scientists (Covid.cd2h.org/duas) who have contributed to the on-going development of this community resource (17). We would like to thank the National Community Engagement Group (NCEG), all patient, caregiver and community Representatives, and all the participants enrolled in the RECOVER Initiative.

The N3C data transfer to NCATS is performed under a Johns Hopkins University Reliance Protocol # IRB00249128 or individual site agreements with NIH. The N3C Data Enclave is managed under the authority of the NIH; information can be found at https://ncats.nih.gov/n3c/resources.

We gratefully acknowledge the following core contributors to N3C:

Adam B. Wilcox, Adam M. Lee, Alexis Graves, Alfred (Jerrod) Anzalone, Amin Manna, Amit Saha, Amy Olex, Andrea Zhou, Andrew E. Williams, Andrew Southerland, Andrew T. Girvin, Anita Walden, Anjali A. Sharathkumar, Benjamin Amor, Benjamin Bates, Brian Hendricks, Brijesh Patel, Caleb Alexander, Carolyn Bramante, Cavin Ward-Caviness, Charisse Madlock-Brown, Christine Suver, Christopher Chute, Christopher Dillon, Chunlei Wu, Clare Schmitt, Cliff Takemoto, Dan Housman, Davera Gabriel, David A. Eichmann, Diego Mazzotti, Don Brown, Eilis Boudreau, Elaine Hill, Elizabeth Zampino, Emily Carlson Marti, Emily R. Pfaff, Evan French, Farrukh M Koraishy, Federico Mariona, Fred Prior, George Sokos, Greg Martin, Harold Lehmann, Heidi Spratt, Hemalkumar Mehta, Hongfang Liu, Hythem Sidky, J.W. Awori Hayanga, Jami Pincavitch, Jaylyn Clark, Jeremy Richard Harper, Jessica Islam, Jin Ge, Joel Gagnier, Joel H. Saltz, Joel Saltz, Johanna Loomba, John Buse, Jomol Mathew, Joni L. Rutter, Julie A. McMurry, Justin Guinney, Justin Starren, Karen Crowley, Katie Rebecca Bradwell, Kellie M. Walters, Ken Wilkins, Kenneth R. Gersing, Kenrick Dwain Cato, Kimberly Murray, Kristin Kostka, Lavance Northington, Lee Allan Pyles, Leonie Misquitta, Lesley Cottrell, Lili Portilla, Mariam Deacy, Mark M. Bissell, Marshall Clark, Mary Emmett, Mary Morrison Saltz, Matvey B. Palchuk, Melissa A. Haendel, Meredith Adams, Meredith Temple-O’Connor, Michael G. Kurilla, Michele Morris, Nabeel Qureshi, Nasia Safdar, Nicole Garbarini, Noha Sharafeldin, Ofer Sadan, Patricia A. Francis, Penny Wung Burgoon, Peter Robinson, Philip R.O. Payne, Rafael Fuentes, Randeep Jawa, Rebecca Erwin-Cohen, Rena Patel, Richard A. Moffitt, Richard L. Zhu, Rishi Kamaleswaran, Robert Hurley, Robert T. Miller, Saiju Pyarajan, Sam G. Michael, Samuel Bozzette, Sandeep Mallipattu, Satyanarayana Vedula, Scott Chapman, Shawn T. O’Neil, Soko Setoguchi, Stephanie S. Hong, Steve Johnson, Tellen D. Bennett, Tiffany Callahan, Umit Topaloglu, Usman Sheikh, Valery Gordon, Vignesh Subbian, Warren A. Kibbe, Wenndy Hernandez, Will Beasley, Will Cooper, William Hillegass, Xiaohan Tanner Zhang. Details of contributions available at Covid.cd2h.org/core-contributors

The following institutions whose data is released or pending:

Available: Advocate Health Care Network — UL1TR002389: The Institute for Translational Medicine (ITM) • Boston University Medical Campus — UL1TR001430: Boston University Clinical and Translational Science Institute • Brown University — U54GM115677: Advance Clinical Translational Research (Advance-CTR) • Carilion Clinic — UL1TR003015: iTHRIV Integrated Translational health Research Institute of Virginia • Charleston Area Medical Center — U54GM104942: West Virginia Clinical and Translational Science Institute (WVCTSI) • Children’s Hospital Colorado — UL1TR002535: Colorado Clinical and Translational Sciences Institute • Columbia University Irving Medical Center — UL1TR001873: Irving Institute for Clinical and Translational Research • Duke University — UL1TR002553: Duke Clinical and Translational Science Institute • George Washington Children’s Research Institute — UL1TR001876: Clinical and Translational Science Institute at Children’s National (CTSA-CN) • George Washington University — UL1TR001876: Clinical and Translational Science Institute at Children’s National (CTSA-CN) • Indiana University School of Medicine — UL1TR002529: Indiana Clinical and Translational Science Institute • Johns Hopkins University — UL1TR003098: Johns Hopkins Institute for Clinical and Translational Research • Loyola Medicine — Loyola University Medical Center • Loyola University Medical Center — UL1TR002389: The Institute for Translational Medicine (ITM) • Maine Medical Center — U54GM115516: Northern New England Clinical & Translational Research (NNE-CTR) Network • Massachusetts General Brigham — UL1TR002541: Harvard Catalyst • Mayo Clinic Rochester — UL1TR002377: Mayo Clinic Center for Clinical and Translational Science (CCaTS) • Medical University of South Carolina — UL1TR001450: South Carolina Clinical & Translational Research Institute (SCTR) • Montefiore Medical Center — UL1TR002556: Institute for Clinical and Translational Research at Einstein and Montefiore • Nemours U54GM104941: Delaware CTR ACCEL Program • NorthShore University HealthSystem — UL1TR002389: The Institute for Translational Medicine (ITM) • Northwestern University at Chicago — UL1TR001422: Northwestern University Clinical and Translational Science Institute (NUCATS) • OCHIN — INV-018455: Bill and Melinda Gates Foundation grant to Sage Bionetworks • Oregon Health & Science University — UL1TR002369: Oregon Clinical and Translational Research Institute • Penn State Health Milton S. Hershey Medical Center — UL1TR002014: Penn State Clinical and Translational Science Institute • Rush University Medical Center — UL1TR002389: The Institute for Translational Medicine (ITM) • Rutgers, The State University of New Jersey UL1TR003017: New Jersey Alliance for Clinical and Translational Science • Stony Brook University — U24TR002306 • The Ohio State University — UL1TR002733: Center for Clinical and Translational Science • The State University of New York at Buffalo — UL1TR001412: Clinical and Translational Science Institute • The University of Chicago — UL1TR002389: The Institute for Translational Medicine (ITM) • The University of Iowa — UL1TR002537: Institute for Clinical and Translational Science • The University of Miami Leonard M. Miller School of Medicine — UL1TR002736: University of Miami Clinical and Translational Science Institute • The University of Michigan at Ann Arbor — UL1TR002240: Michigan Institute for Clinical and Health Research • The University of Texas Health Science Center at Houston — UL1TR003167: Center for Clinical and Translational Sciences (CCTS) • The University of Texas Medical Branch at Galveston — UL1TR001439: The Institute for Translational Sciences • The University of Utah — UL1TR002538: Uhealth Center for Clinical and Translational Science • Tufts Medical Center — UL1TR002544: Tufts Clinical and Translational Science Institute • Tulane University — UL1TR003096: Center for Clinical and Translational Science • University Medical Center New Orleans — U54GM104940: Louisiana Clinical and Translational Science (LA CaTS) Center • University of Alabama at Birmingham — UL1TR003096: Center for Clinical and Translational Science • University of Arkansas for Medical Sciences — UL1TR003107: UAMS Translational Research Institute • University of Cincinnati — UL1TR001425: Center for Clinical and Translational Science and Training • University of Colorado Denver, Anschutz Medical Campus — UL1TR002535: Colorado Clinical and Translational Sciences Institute • University of Illinois at Chicago — UL1TR002003: UIC Center for Clinical and Translational Science • University of Kansas Medical Center — UL1TR002366: Frontiers: University of Kansas Clinical and Translational Science Institute • University of Kentucky — UL1TR001998: UK Center for Clinical and Translational Science • University of Massachusetts Medical School Worcester — UL1TR001453: The UMass Center for Clinical and Translational Science (UMCCTS) • University of Minnesota — UL1TR002494: Clinical and Translational Science Institute • University of Mississippi Medical Center — U54GM115428: Mississippi Center for Clinical and Translational Research (CCTR) • University of Nebraska Medical Center — U54GM115458: Great Plains IDeA-Clinical & Translational Research • University of North Carolina at Chapel Hill — UL1TR002489: North Carolina Translational and Clinical Science Institute • University of Oklahoma Health Sciences Center — U54GM104938: Oklahoma Clinical and Translational Science Institute (OCTSI) • University of Rochester — UL1TR002001: UR Clinical & Translational Science Institute • University of Southern California — UL1TR001855: The Southern California Clinical and Translational Science Institute (SC CTSI) • University of Vermont — U54GM115516: Northern New England Clinical & Translational Research (NNE-CTR) Network • University of Virginia — UL1TR003015: iTHRIV Integrated Translational health Research Institute of Virginia • University of Washington — UL1TR002319: Institute of Translational Health Sciences • University of Wisconsin-Madison — UL1TR002373: UW Institute for Clinical and Translational Research • Vanderbilt University Medical Center — UL1TR002243: Vanderbilt Institute for Clinical and Translational Research • Virginia Commonwealth University — UL1TR002649: C. Kenneth and Dianne Wright Center for Clinical and Translational Research • Wake Forest University Health Sciences — UL1TR001420: Wake Forest Clinical and Translational Science Institute • Washington University in St. Louis — UL1TR002345: Institute of Clinical and Translational Sciences • Weill Medical College of Cornell University — UL1TR002384: Weill Cornell Medicine Clinical and Translational Science Center • West Virginia University — U54GM104942: West Virginia Clinical and Translational Science Institute (WVCTSI) Submitted: Icahn School of Medicine at Mount Sinai — UL1TR001433: ConduITS Institute for Translational Sciences • The University of Texas Health Science Center at Tyler — UL1TR003167: Center for Clinical and Translational Sciences (CCTS) • University of California, Davis — UL1TR001860: UCDavis Health Clinical and Translational Science Center • University of California, Irvine — UL1TR001414: The UC Irvine Institute for Clinical and Translational Science (ICTS) • University of California, Los Angeles — UL1TR001881: UCLA Clinical Translational Science Institute • University of California, San Diego — UL1TR001442: Altman Clinical and Translational Research Institute • University of California, San Francisco — UL1TR001872: UCSF Clinical and Translational Science Institute Pending: Arkansas Children’s Hospital — UL1TR003107: UAMS Translational Research Institute • Baylor College of Medicine — None (Voluntary) • Children’s Hospital of Philadelphia — UL1TR001878: Institute for Translational Medicine and Therapeutics • Cincinnati Children’s Hospital Medical Center — UL1TR001425: Center for Clinical and Translational Science and Training • Emory University — UL1TR002378: Georgia Clinical and Translational Science Alliance • HonorHealth — None (Voluntary) • Loyola University Chicago — UL1TR002389: The Institute for Translational Medicine (ITM) • Medical College of Wisconsin — UL1TR001436: Clinical and Translational Science Institute of Southeast Wisconsin • MedStar Health Research Institute — UL1TR001409: The Georgetown-Howard Universities Center for Clinical and Translational Science (GHUCCTS) • MetroHealth — None (Voluntary) • Montana State University — U54GM115371: American Indian/Alaska Native CTR • NYU Langone Medical Center — UL1TR001445: Langone Health’s Clinical and Translational Science Institute • Ochsner Medical Center — U54GM104940: Louisiana Clinical and Translational Science (LA CaTS) Center • Regenstrief Institute — UL1TR002529: Indiana Clinical and Translational Science Institute • Sanford Research — None (Voluntary) • Stanford University — UL1TR003142: Spectrum: The Stanford Center for Clinical and Translational Research and Education • The Rockefeller University — UL1TR001866: Center for Clinical and Translational Science • The Scripps Research Institute — UL1TR002550: Scripps Research Translational Institute • University of Florida — UL1TR001427: UF Clinical and Translational Science Institute • University of New Mexico Health Sciences Center — UL1TR001449: University of New Mexico Clinical and Translational Science Center • University of Texas Health Science Center at San Antonio — UL1TR002645: Institute for Integration of Medicine and Science • Yale New Haven Hospital — UL1TR001863: Yale Center for Clinical Investigation

