## Supplementary Table 1; Supplementary Table 2 for "Are fewer cases of diabetes mellitus diagnosed in the months after SARS-CoV-2 infection?"

**Supplementary Appendix**

**Supplementary Table 1: Proportion of individuals in the electronic health record database with type 2 diabetes mellitus whose diabetes was diagnosed after September 2019. These data are also represented in for Figure 1.**

| Day relative to infection | -180  to  -150 | -149  to  -120 | -119  to  -90 | -89  to  -60 | -59  to  -30 | -30  to  -8 | -8  to  +30 | 31  to  60 | 61  to  90 | 91  to  120 | 121  to  150 | 151  to  180 | 180  to  210 | 211  to  240 | 241  to  270 | 271  to  300 | 301  to  330 | 331  to  360 |
| --- | --- | --- | --- | --- | --- | --- | --- | --- | --- | --- | --- | --- | --- | --- | --- | --- | --- | --- |
| 3-Mar-20 | 0.0038 | 0.0028 | 0.0022 | 0.0024 | 0.0027 | 0.0025 | 0.0263 | 0.0017 | 0.0013 | 0.0015 | 0.0015 | 0.0015 | 0.0013 | 0.0014 | 0.0015 | 0.0011 |  | 0.0017 |
| 4-Apr-20 | 0.0038 | 0.0026 | 0.0026 | 0.0033 | 0.0027 | 0.0018 | 0.0225 | 0.0018 | 0.0015 | 0.0012 | 0.0014 | 0.0016 | 0.0014 | 0.0013 | 0.0011 | 0.0014 | 0.0013 | 0.0013 |
| 5-May-20 | 0.0033 | 0.0024 | 0.0033 | 0.0026 | 0.0016 | 0.0014 | 0.0153 | 0.0019 | 0.0014 | 0.0015 | 0.0016 | 0.0019 | 0.0015 | 0.0011 | 0.0013 | 0.0014 | 0.0014 | 0.0012 |
| 6-Jun-20 | 0.0021 | 0.0026 | 0.0025 | 0.0019 | 0.0017 | 0.0018 | 0.0141 | 0.0020 | 0.0013 | 0.0016 | 0.0017 | 0.0014 | 0.0010 | 0.0012 | 0.0011 | 0.0012 | 0.0013 | 0.0012 |
| 7-Jul-20 | 0.0024 | 0.0018 | 0.0016 | 0.0018 | 0.0013 | 0.0013 | 0.0115 | 0.0017 | 0.0014 | 0.0014 | 0.0008 | 0.0011 | 0.0012 | 0.0011 | 0.0012 | 0.0011 | 0.0009 | 0.0007 |
| 8-Aug-20 | 0.0019 | 0.0012 | 0.0018 | 0.0018 | 0.0021 | 0.0019 | 0.0124 | 0.0019 | 0.0017 | 0.0013 | 0.0010 | 0.0012 | 0.0014 | 0.0012 | 0.0011 | 0.0009 | 0.0010 | 0.0011 |
| 9-Sep-20 | 0.0012 | 0.0014 | 0.0016 | 0.0023 | 0.0017 | 0.0018 | 0.0110 | 0.0018 | 0.0013 | 0.0010 | 0.0010 | 0.0012 | 0.0009 | 0.0010 | 0.0013 | 0.0011 | 0.0011 | 0.0009 |
| 10-Oct-20 | 0.0013 | 0.0017 | 0.0015 | 0.0016 | 0.0017 | 0.0015 | 0.0099 | 0.0017 | 0.0012 | 0.0014 | 0.0012 | 0.0013 | 0.0012 | 0.0010 | 0.0011 | 0.0009 | 0.0010 | 0.0008 |
| 11-Nov-20 | 0.0017 | 0.0015 | 0.0016 | 0.0015 | 0.0017 | 0.0013 | 0.0084 | 0.0016 | 0.0014 | 0.0013 | 0.0012 | 0.0012 | 0.0010 | 0.0009 | 0.0010 | 0.0010 | 0.0010 | 0.0011 |
| 12-Dec-20 | 0.0018 | 0.0017 | 0.0018 | 0.0020 | 0.0024 | 0.0014 | 0.0099 | 0.0020 | 0.0017 | 0.0013 | 0.0013 | 0.0012 | 0.0011 | 0.0010 | 0.0010 | 0.0010 | 0.0010 | 0.0012 |
| 13-Jan-21 | 0.0021 | 0.0020 | 0.0022 | 0.0024 | 0.0018 | 0.0012 | 0.0113 | 0.0022 | 0.0015 | 0.0013 | 0.0011 | 0.0012 | 0.0012 | 0.0010 | 0.0010 | 0.0012 | 0.0011 | 0.0010 |
| 14-Feb-21 | 0.0022 | 0.0020 | 0.0024 | 0.0016 | 0.0019 | 0.0015 | 0.0147 | 0.0022 | 0.0016 | 0.0015 | 0.0015 | 0.0013 | 0.0011 | 0.0008 | 0.0010 | 0.0012 | 0.0010 | 0.0009 |
| 15-Mar-21 | 0.0022 | 0.0023 | 0.0016 | 0.0015 | 0.0021 | 0.0019 | 0.0138 | 0.0019 | 0.0016 | 0.0012 | 0.0011 | 0.0010 | 0.0009 | 0.0012 | 0.0011 | 0.0010 | 0.0009 | 0.0007 |
| 16-Apr-21 | 0.0018 | 0.0016 | 0.0016 | 0.0015 | 0.0022 | 0.0016 | 0.0123 | 0.0017 | 0.0012 | 0.0012 | 0.0010 | 0.0009 | 0.0010 | 0.0007 | 0.0008 | 0.0006 | 0.0011 | 0.0011 |
| 17-May-21 | 0.0017 | 0.0018 | 0.0021 | 0.0021 | 0.0018 | 0.0018 | 0.0152 | 0.0020 | 0.0014 | 0.0013 | 0.0012 | 0.0010 | 0.0012 | 0.0011 | 0.0010 | 0.0009 | 0.0011 | 0.0007 |
| 18-Jun-21 | 0.0018 | 0.0020 | 0.0024 | 0.0018 | 0.0021 | 0.0024 | 0.0142 | 0.0015 |  |  | 0.0015 |  | 0.0012 |  | 0.0013 | 0.0012 |  |  |
| 19-Jul-21 | 0.0020 | 0.0017 | 0.0015 | 0.0019 | 0.0017 | 0.0011 | 0.0122 | 0.0013 | 0.0010 | 0.0013 | 0.0009 | 0.0006 | 0.0010 | 0.0006 | 0.0007 | 0.0006 |  |  |
| 20-Aug-21 | 0.0016 | 0.0017 | 0.0014 | 0.0016 | 0.0013 | 0.0011 | 0.0105 | 0.0011 | 0.0010 | 0.0011 | 0.0010 | 0.0009 | 0.0010 | 0.0010 | 0.0005 | 0.0003 |  |  |
| 21-Sep-21 | 0.0016 | 0.0016 | 0.0015 | 0.0016 | 0.0016 | 0.0011 | 0.0109 | 0.0016 | 0.0013 | 0.0011 | 0.0010 | 0.0010 | 0.0013 | 0.0007 | 0.0004 |  |  |  |
| 22-Oct-21 | 0.0018 | 0.0014 | 0.0013 | 0.0016 | 0.0016 | 0.0014 | 0.0111 | 0.0015 | 0.0011 | 0.0012 | 0.0014 | 0.0010 | 0.0007 | 0.0006 |  |  |  |  |
| 23-Nov-21 | 0.0014 | 0.0015 | 0.0013 | 0.0013 | 0.0015 | 0.0013 | 0.0094 | 0.0015 | 0.0011 | 0.0011 | 0.0011 | 0.0005 | 0.0002 |  |  |  |  |  |
| 24-Dec-21 | 0.0016 | 0.0013 | 0.0012 | 0.0015 | 0.0015 | 0.0011 | 0.0075 | 0.0014 | 0.0011 | 0.0010 | 0.0006 | 0.0003 |  |  |  |  |  |  |
| 25-Jan-22 | 0.0015 | 0.0015 | 0.0015 | 0.0016 | 0.0016 | 0.0011 | 0.0063 | 0.0015 | 0.0014 | 0.0007 | 0.0004 | 0.0001 |  |  |  |  |  |  |
| 26-Feb-22 | 0.0021 | 0.0020 | 0.0023 | 0.0021 | 0.0020 | 0.0016 | 0.0113 | 0.0017 | 0.0012 | 0.0008 | 0.0003 |  |  |  |  |  |  |  |
| 29-May-22 | 0.0017 | 0.0013 | 0.0017 | 0.0018 | 0.0018 | 0.0015 | 0.0047 |  |  |  |  |  |  |  |  |  |  |  |

Proportion of all persons with a new type 2 diabetes mellitus diagnosis who received that ICD code from 6 months prior to 360 days after SARS-CoV-2 infection, per month of the pandemic.

**Supplementary Table 2: Number of new type 2 diabetes mellitus cases by month of SARS-CoV-2 infection. These data are also represented in Figure 2.**

| **Total count** | Day relative to infection | -180  to  -50 | -149  to  -120 | -119  to  -90 | -89  to  -60 | -59  to  -30 | -30  to  -8 | -8  to  +30 | 31  to  60 | 61  to  90 | 91  to  120 | 121  to  150 | 151  to  180 | 180  to  210 | 211  to  240 | 241  to  270 | 271  to  300 | 301  to  330 | 331  to  360 |
| --- | --- | --- | --- | --- | --- | --- | --- | --- | --- | --- | --- | --- | --- | --- | --- | --- | --- | --- | --- |
| 21988 | 3-Mar-20 | 83 | 61 | 48 | 52 | 60 | 56 | 579 | 38 | 29 | 32 | 34 | 34 | 29 | 31 | 33 | 25 | <20 | 37 |
| 63018 | 4-Apr-20 | 239 | 166 | 163 | 207 | 168 | 113 | 1421 | 111 | 95 | 76 | 87 | 98 | 91 | 82 | 70 | 87 | 83 | 80 |
| 49149 | 5-May-20 | 163 | 118 | 163 | 128 | 77 | 67 | 754 | 92 | 71 | 76 | 77 | 92 | 73 | 52 | 62 | 71 | 68 | 57 |
| 41683 | 6-Jun-20 | 88 | 110 | 103 | 78 | 72 | 73 | 586 | 82 | 55 | 65 | 70 | 59 | 40 | 48 | 46 | 49 | 53 | 50 |
| 66264 | 7-Jul-20 | 160 | 118 | 105 | 116 | 88 | 84 | 760 | 112 | 95 | 92 | 56 | 74 | 79 | 70 | 82 | 74 | 58 | 44 |
| 59857 | 8-Aug-20 | 113 | 73 | 106 | 105 | 128 | 114 | 741 | 115 | 101 | 77 | 60 | 71 | 85 | 74 | 63 | 51 | 62 | 63 |
| 55507 | 9-Sep-20 | 65 | 79 | 89 | 126 | 95 | 98 | 611 | 98 | 71 | 55 | 53 | 68 | 51 | 56 | 72 | 61 | 59 | 48 |
| 106177 | 10-Oct-20 | 138 | 180 | 162 | 170 | 184 | 162 | 1048 | 181 | 131 | 148 | 129 | 140 | 129 | 107 | 118 | 96 | 101 | 85 |
| 244876 | 11-Nov-20 | 409 | 378 | 390 | 378 | 420 | 328 | 2048 | 380 | 345 | 324 | 304 | 285 | 255 | 230 | 245 | 241 | 244 | 270 |
| 253434 | 12-Dec-20 | 468 | 434 | 446 | 516 | 597 | 354 | 2518 | 498 | 427 | 335 | 342 | 300 | 278 | 260 | 256 | 251 | 264 | 303 |
| 177844 | 13-Jan-21 | 378 | 353 | 386 | 425 | 315 | 220 | 2013 | 392 | 270 | 240 | 193 | 215 | 219 | 170 | 178 | 205 | 198 | 178 |
| 72152 | 14-Feb-21 | 161 | 144 | 176 | 116 | 134 | 106 | 1062 | 159 | 113 | 110 | 107 | 94 | 81 | 60 | 71 | 83 | 71 | 65 |
| 59866 | 15-Mar-21 | 132 | 137 | 93 | 88 | 123 | 115 | 828 | 116 | 93 | 73 | 64 | 62 | 56 | 69 | 64 | 59 | 53 | 43 |
| 63475 | 16-Apr-21 | 115 | 99 | 100 | 96 | 137 | 100 | 783 | 109 | 76 | 74 | 61 | 58 | 61 | 46 | 51 | 37 | 69 | 70 |
| 37838 | 17-May-21 | 65 | 67 | 79 | 79 | 69 | 70 | 576 | 74 | 54 | 48 | 45 | 37 | 47 | 40 | 36 | 33 | 40 | 25 |
| 19748 | 18-Jun-21 | 35 | 39 | 47 | 35 | 42 | 48 | 280 | 29 | <20 | <20 | 30 | <20 | 23 | <20 | 26 | 24 | <20 | <20 |
| 39539 | 19-Jul-21 | 79 | 68 | 59 | 74 | 66 | 42 | 481 | 50 | 40 | 53 | 36 | 25 | 38 | 23 | 26 | 24 | <20 | <20 |
| 126287 | 20-Aug-21 | 197 | 219 | 183 | 205 | 170 | 142 | 1325 | 142 | 131 | 133 | 127 | 117 | 128 | 131 | 59 | 37 | <20 | <20 |
| 120129 | 21-Sep-21 | 192 | 191 | 175 | 187 | 194 | 130 | 1304 | 198 | 154 | 129 | 119 | 117 | 155 | 87 | 43 | <20 | <20 | <20 |
| 75960 | 22-Oct-21 | 139 | 110 | 99 | 118 | 118 | 105 | 845 | 117 | 85 | 89 | 109 | 78 | 51 | 42 | <20 | <20 | <20 | <20 |
| 97887 | 23-Nov-21 | 135 | 144 | 129 | 132 | 142 | 131 | 920 | 149 | 107 | 111 | 108 | 48 | 24 | <20 | <20 | <20 | <20 | <20 |
| 224781 | 24-Dec-21 | 350 | 298 | 277 | 342 | 330 | 248 | 1681 | 321 | 249 | 218 | 126 | 64 | <20 | <20 | <20 | <20 | <20 | <20 |
| 424738 | 25-Jan-22 | 649 | 616 | 639 | 679 | 668 | 462 | 2655 | 651 | 594 | 291 | 150 | 30 | <20 | <20 | <20 | <20 | <20 | <20 |
| 76582 | 26-Feb-22 | 157 | 151 | 177 | 162 | 154 | 125 | 864 | 132 | 91 | 58 | 21 | <20 | <20 | <20 | <20 | <20 | <20 | <20 |
| 24794 | 27-Mar-22 | 55 | 48 | 40 | 40 | 56 | 49 | 282 | 28 | <20 | <20 | <20 | <20 | <20 | <20 | <20 | <20 | <20 | <20 |
| 35539 | 28-Apr-22 | 56 | 67 | 50 | 62 | 62 | 50 | 190 | <20 | <20 | <20 | <20 | <20 | <20 | <20 | <20 | <20 | <20 | <20 |
| 54951 | 29-May-22 | 92 | 69 | 92 | 99 | 101 | 85 | 256 | <20 | <20 | <20 | <20 | <20 | <20 | <20 | <20 | <20 | <20 | <20 |
| 31044 | 30-Jun-22 | 57 | 53 | 66 | 76 | 67 | 40 | 152 | <20 | <20 | <20 | <20 | <20 | <20 | <20 | <20 | <20 | <20 | <20 |

Total count is the total number of people in the N3C Data Enclave with COVID-19 and a record of type 2 diabetes who have at least one clinical encounter in the database greater than 6 months prior to COVID-19 diagnosis. Each row is the number of new type 2 diabetes mellitus cases reported by month of COVID-19 diagnosis. Each column is the number of days from the index date. Day 1 is the first date of COVID-19 diagnosis by either lab result or problem list.
